# Transmission characteristics of the COVID-19 outbreak in China: a study driven by data

**DOI:** 10.1101/2020.02.26.20028431

**Authors:** Meili Li, Pian Chen, Qianqian Yuan, Baojun Song, Junling Ma

## Abstract

The COVID-19 outbreak has been a serious public health threat worldwide. We use individually documented case descriptions of COVID-19 from China (excluding Hubei Province) to estimate the distributions of the generation time, incubation period, and periods from symptom onset to isolation and to diagnosis. The recommended 14-day quarantine period may lead to a 6.7% failure for quarantine. We recommend a 22-day quarantine period. The mean generation time is 3.3 days and the mean incubation period is 7.2 days. It took 3.7 days to isolate and 6.6 days to diagnose a patient after his/her symptom onset. Patients may become infectious on average 3.9 days before showing major symptoms. This makes contact tracing and quarantine ineffective. The basic reproduction number is estimated to be 1.54 with contact tracing, quarantine and isolation, mostly driven by super spreaders.

---

The COVID-19 outbreak in China has become a major global public health concern (1-3). Many estimates of the scope and magnitude of the outbreak have been quickly published (4-7). The basic reproduction number, which is the average number of secondary infections caused by an average infectious individual during entire infectious period in a susceptible population, has received much attention, as the epidemic can take off if ℛ_0_ > 1. The basic reproduction number ℛ_0_ for the novel coronavirus has been estimated to be between 2.2 and 6.47 (8-12). Unfortunately, the most important disease characteristic parameters such as the generation time (the time between being infected and infecting others), the incubation period, and the periods from symptom onset to diagnosis and isolation, are still unknown. The published studies used educated guesses, many of such guesses were based on SARS epidemics. However, these studies also suggest that COVID-19 outbreak has different characteristics than SARS, such as possible transmission before showing major symptoms, and possibly higher transmission rates. Thus, SARS epidemics may not be a reliable source of information for COVID-19.

We aim to estimate the distributions of important periods related to the disease progression and transmission of SARS-CoV-2, including the generation time, the incubation period, the periods from symptom onset to isolation and diagnosis. We also aim to estimate the basic reproduction number for COVID-19 in Chinese provinces excluding Hubei (where Wuhan is the provincial capital), because these provinces implemented strong contact tracing, quarantine, and isolation control strategies. The outbreak patterns in these provinces thus may provide crucial information for the control of COVID-19 outbreaks in other countries.

Each province except Hubei officially published the daily update of case descriptions of COVID-19. We use these individually documented case descriptions to conduct our research. The documented cases and their data sources are tabulated in Supplementary Material S1. Some of these descriptions are well documented with important epidemiological information including both the contact information and dates such as symptom onset, isolation and diagnosis. We fit the data to a gamma distribution and a log-normal distribution (and also an exponential distribution for the period from symptom onset to isolation) using Markov Chain Monte Carlo (MCMC), and select the one with the smallest Deviance Information Criterion (DIC). The case descriptions also allow us to construct graph of transmissions to estimate the basic reproduction number ℛ_0_. The details of the estimation procedures are described in Supplementary Material S2.

The distribution of the period from symptom onset to quarantine (shown as negative periods) or isolation (shown as positive periods) is shown in Figure 1. Voluntary quarantine at home is not counted here, because they may still cause infection among family members, through whom the infection can be leaked outside households. Since the lockdown of Wuhan on January 23, 2020 may significantly increase the awareness among the public, and subsequently change individual behaviors, we also compare the fitted distribution for patients who showed symptom before January 23, and those who showed symptom after January 23. Overall, the period from symptom onset to isolation follows a gamma distribution with a mean 3.7 (95% confidence limits are 3.5-3.9) days and a variance 16.1 (13.8, 18.7), corresponding to a shape parameter 0.85 (0.78, 0.93) and a scale parameter 4.35 (3.92, 4.81). The period for patients showing symptom before January 23 follows a gamma distribution with a mean 5.2 (4.8, 5.6) days and a variance 18.6 (15.8, 19.9), which correspond to a shape parameter 1.46 (1.26, 1.67) and a scale parameter 3.57 (3.15, 3.92). The period for those showing symptom after January 23 follows a gamma distribution with a mean 3.1 (2.9, 3.4) days and a variance 12.9 (10.8, 15.5), corresponding to a shape parameter 0.77 (0.69, 0.86) and a scale parameter 4.10 (3.60, 4.67). Even though the earliest quarantine date is January 21, 2020, only 2.5% patients have a negative period (which means that they are quarantined before they showed symptom). Thus, quarantine has only a small effect in controlling the outbreak.

**Figure 1.**
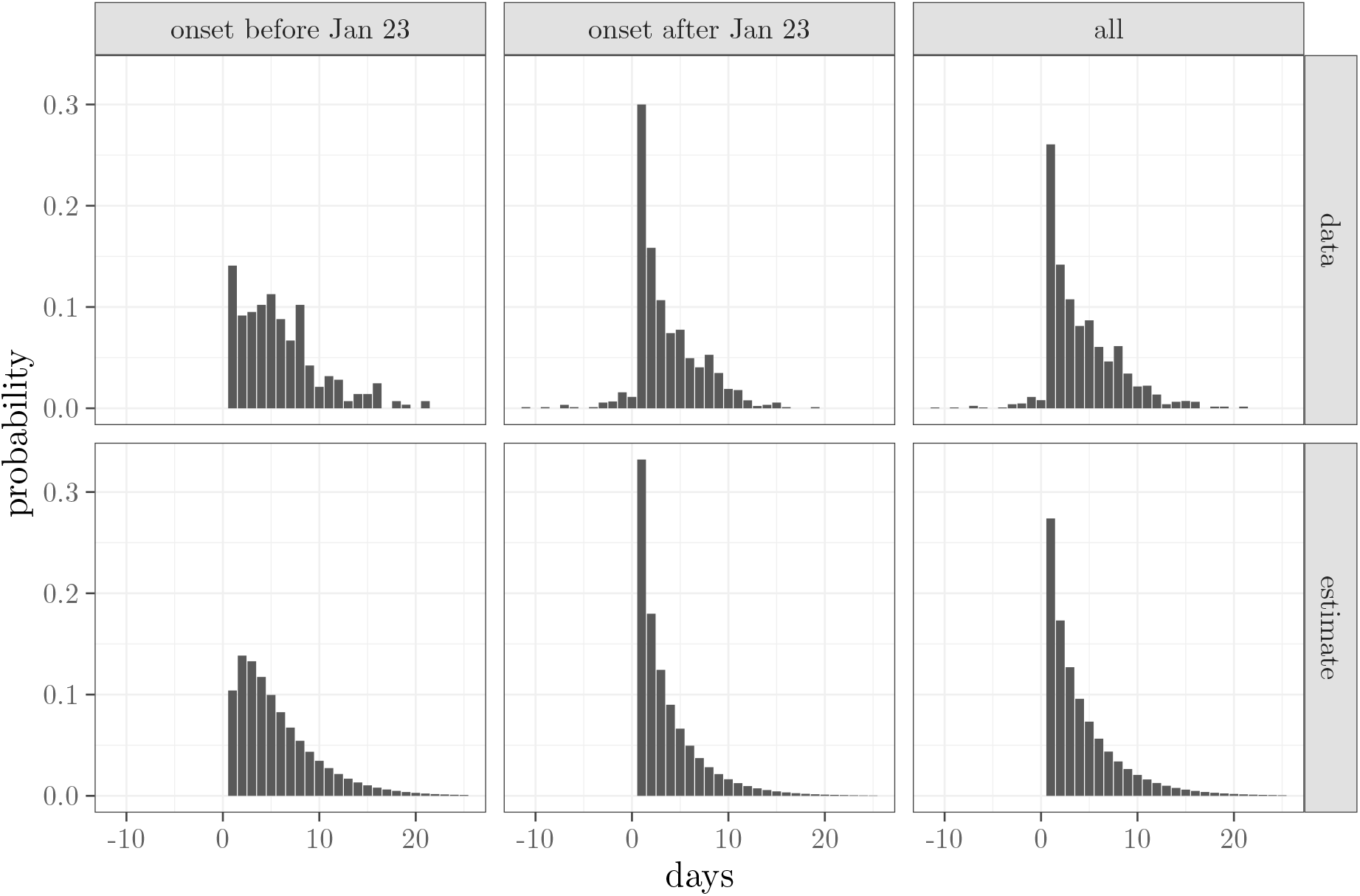
The upper panels show he observed distributions of the period from symptom onset to quarantine (negative periods) or isolation (positive periods). The lower panels show the best fit distributions of the period from symptom onset to isolation calculated from the mean of MCMC samples. The quarantined patients were isolated on the same day of symptom onset. There is a clear difference between individuals who showed symptom before and after January 23, 2020,

The observed and estimated distributions of the period from symptom onset to diagnosis are shown in Figure 2. Overall, the best fit distribution is gamma distributed with a mean 6.6 (6.5, 6.8) days, and a variance 15.9 (14.7, 17.2), corresponding to a shape parameter 2.77 (2.61, 2.94) and a scale parameter 2.40 (2.25, 2.55). For those showing symptom before January 23, the period is estimated to be gamma distributed with a mean 9.3 (8.9, 9.6) days, and a variance 18.5 (16.3, 19.9), corresponding to a shape parameter 4.67 (4.26, 5.18) and a scale parameter 1.99 (1.79, 2.15). For those showing symptom after January 23, the distribution is gamma distributed with a mean 5.5 (5.4, 5.7) days and a variance 10.9 (9.9, 12.0), corresponding to a shape parameter 2.82 (2.61, 3.03) and a scale parameter 1.97 (1.82, 2.13). Diagnosis became much faster after January 23.

**Figure 2.**
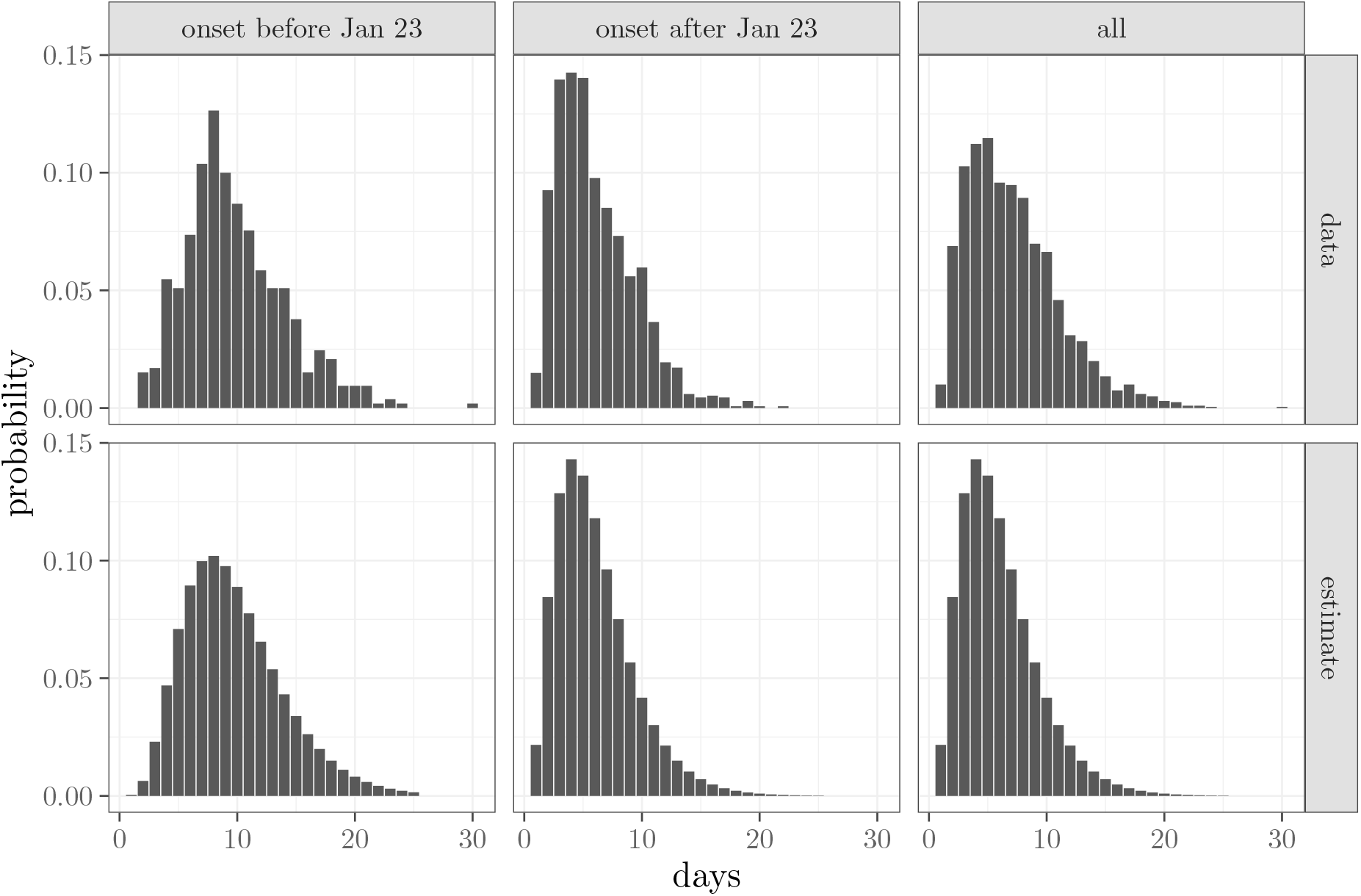
The observed and best fit distributions of the period from symptom onset to diagnosis calculated from the mean of MCMC samples. There is a clear difference between individuals who showed symptom before and after January 23, 2020.

The graph of transmissions is shown in Figure 3. Some patients may have multiple sources of infection. The distribution of the number of secondary infections is shown in Figure 4. Most patients (64.0%) infected one or less individuals. If an individual was infected by one of *n* patients, then the individual only counts as 1/*n* secondary infections of each of the *n* patients. The basic reproduction number can be naively estimated by taking the average of secondary infections over all patients. However, this may be inaccurate because there may be correlations between the number of secondary transmissions of two contacting patients. If individuals with high number of secondary infections are more likely to infect each other, then the average will underestimate ℛ_0_. To properly estimate ℛ_0_, we construct a next generation matrix *N*, in which the (*i, j*) entry is the average number of infectious individuals who made *j* secondary infections caused by an individual who made *i* secondary infections; see (13,14), and Supplementary Material S2 for details. Mathematically,

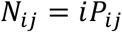

where *P*_*ij*_ is the probability that an individual who infected *j* others was infected by a patient who infected *i* individuals. The basic reproduction number ℛ_0_ is the dominant eigenvalue of *N*. The average number of secondary infections is calculated from data is 1.53. The next generation method gives ℛ_0_ = 1.54. Thus, the correlation of secondary infections has negligible effect on ℛ_0_.

**Figure 3.**
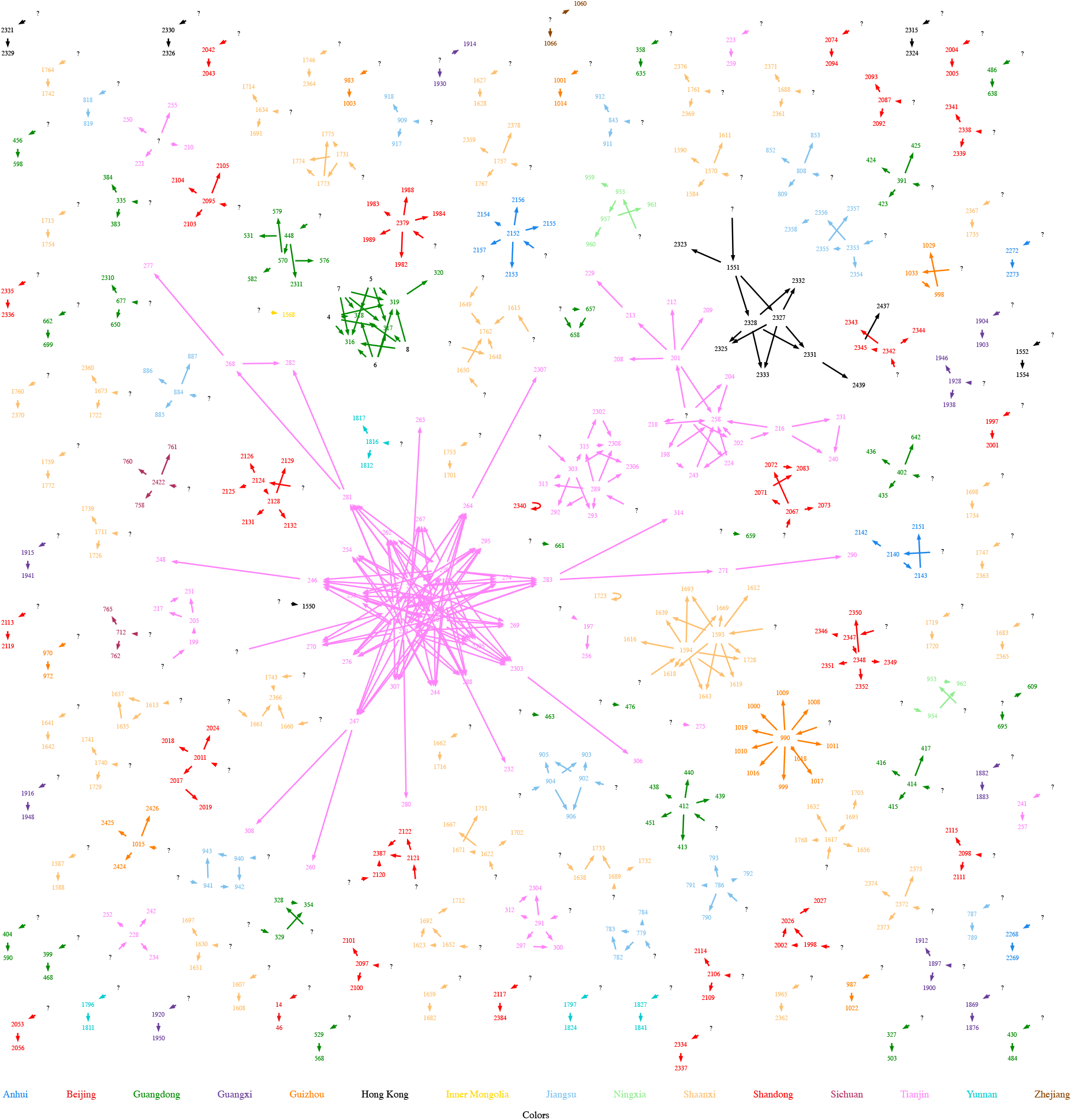
The graph of transmissions for individually documented cases with a contact tracing information. The nodes are patients, and edges show possible transmission. The nodes and edges are colored by province. Patient information, including their labels and provinces, are listed in Supplementary Material S1.

**Figure 4.**
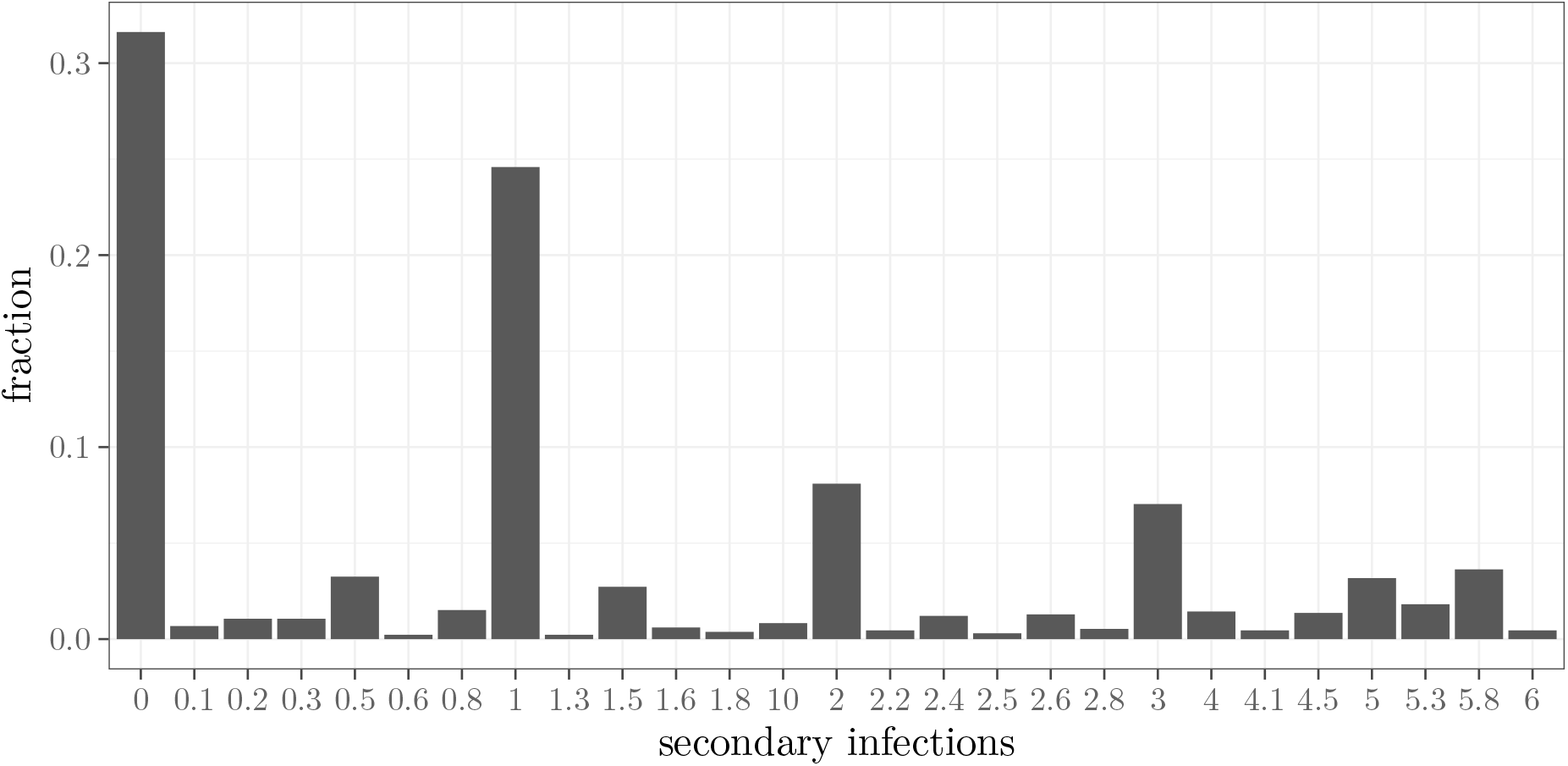
The observed secondary infections caused by a patient. If an individual may be infected by n patients, then the individual counts 1/n as each patient’s secondary infection. The fraction of patients caused one or less secondary infections is 64.0%. The average number of secondary infections is 1.53.

The incubation period distribution is estimated from the nodes in Figure 3 with contact date information. The distribution of the generation time is estimated from the edges in Figure 3 with contact date information for both the source and target nodes. The results are shown in Figure 5. The best fit incubation period distribution is a gamma distribution with a mean 7.2 (6.8, 7.6) days and a variance 16.9 (14.0, 20.2), which correspond to a shape parameter 3.07 (2.62, 3.56) and a scale parameter 2.35 (2.00, 2.75). The generation time is estimated to be gamma distributed with a mean 3.3 (2.3, 4.3) days and a variance 3.1 (1.0, 8.0), corresponding to a shape parameter 4.44 (1.32, 10.02), and a scale parameter 0.95 (0.32, 2.32). Since the mean generation time is smaller than the mean incubation period, on average, a patient may become infectious 3.9 days before showing major symptoms.

**Figure 5.**
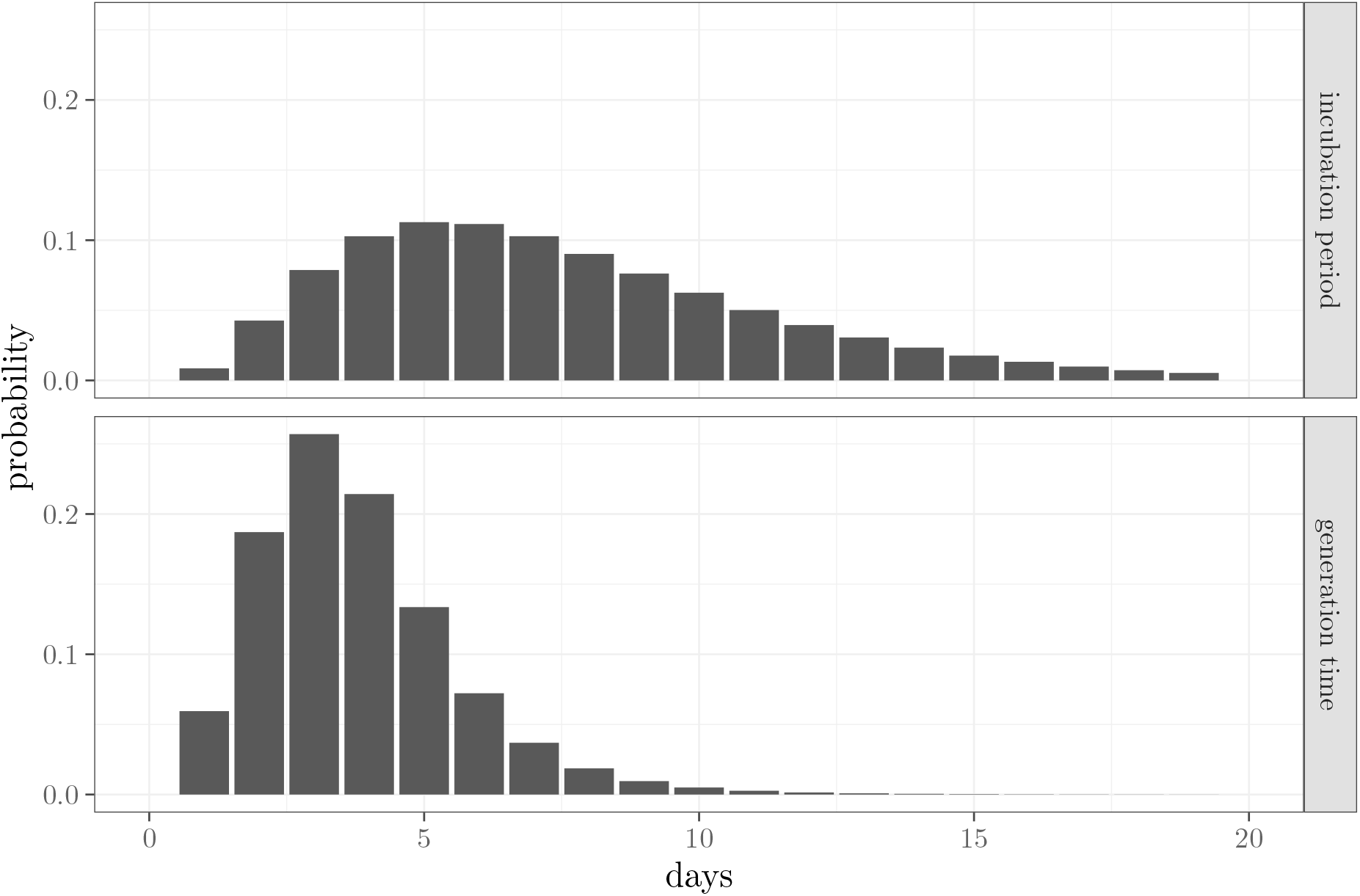
The estimated distributions for the incubation period and the generation time calculated from the mean of the MCMC samples. The mean incubation period is longer than the mean generation time, implying that patients may become infectious before they show major symptoms.

Figure 6 shows the probability distribution of the fraction of individuals whose incubation period is longer than 14, 21, and 22 days, respectively. On average, a quarantine period of 14 days may lead to a failure rate of 6.7%, i.e., 6.7% quarantined patients may show symptom after quarantine. If we aim to control the failure rate of quarantine to be below 1% with 95% confidence, then the quarantine period must be at least 22 days.

**Figure 6.**
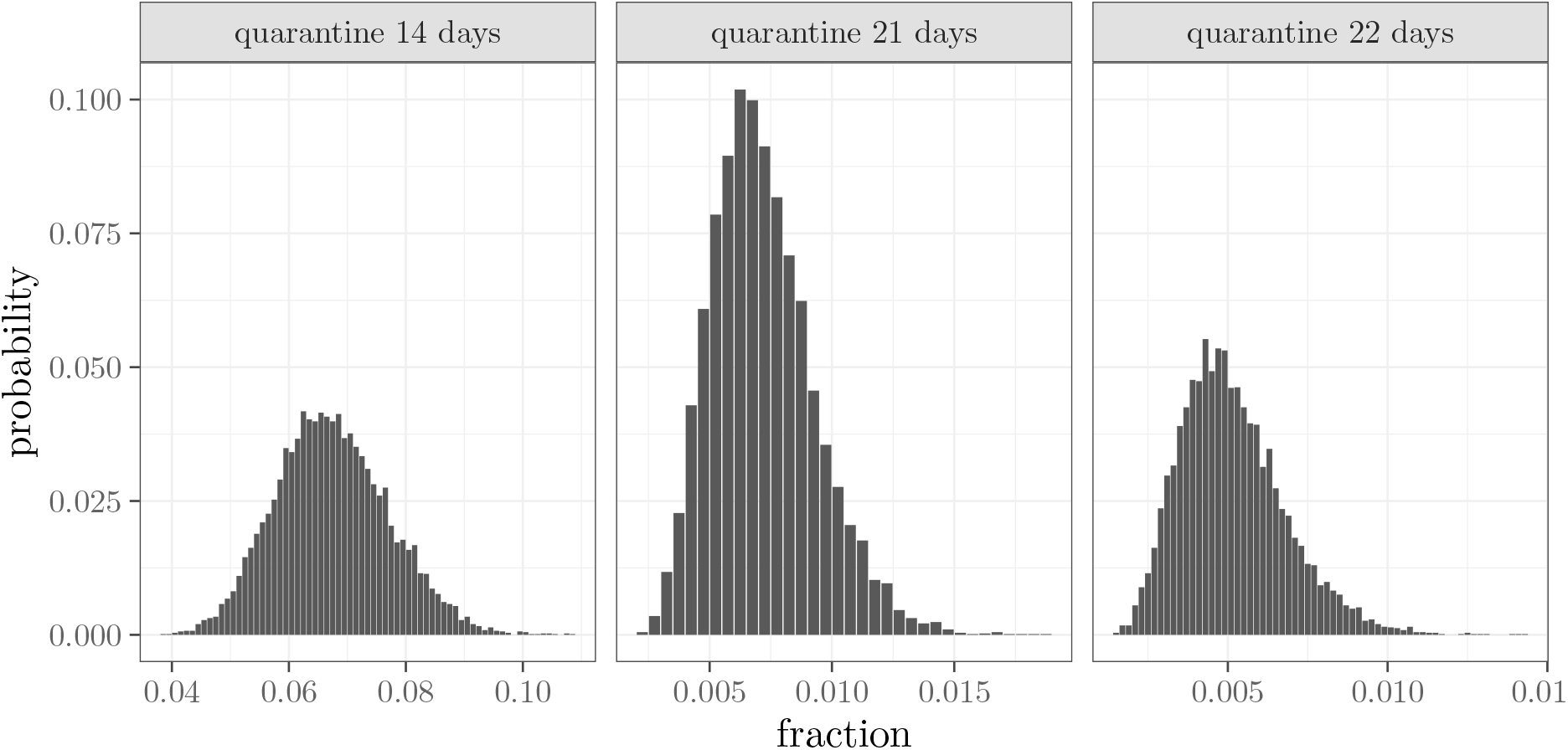
The distributions of the fraction of the patients whose incubation period is longer than 14, 21, and 22 days, represented by the histograms of the probabilities of showing symptoms after the quarantine period calculated from the MCMC samples.

In summary, we estimated the distribution of the generation time, incubation period, and the periods from symptom onset to isolation and to diagnosis for patients in Chinese provinces excluding Hubei. The current recommendation of 14-day quarantine period may be too short, resulting in a 6.7% failure rate. We recommend to increase the quarantine period to 22 days. The periods from symptom onset to isolation and to diagnosis changed significantly for patients who showed symptom before and after the lockdown of Wuhan on January 23, mostly likely driven by behavior change of the general public, and more effective public health control measures after January 23. On average, patients may become infectious 3.9 days before the onset of major symptoms. This, and the 6.6 days delay from symptom onset to diagnosis, severely hinders the effectiveness of contact tracing and quarantine, as evidenced by the 2.5% success rate of quarantine before symptom onset. The basic reproduction number is 1.54 with contact tracing, quarantine and isolation. However, the majority of patients infects no more than one individual, and thus the outbreaks in these provinces are mostly driven by super spreaders. Because transmission can occur before symptom onset, the latent period (from being infected to becoming infectious) and infectious period cannot easily be estimated from case descriptions.

## Data Availability

Data will be made available once it is made into publishable format and validated.

## Acknowledgements

This research is supported by National Natural Science Foundation of China (No. 11771075) (ML), State Scholarship Fund of China (CSC No. 201906635011) (ML), a Fundamental Research Grant for Chinese Universities (ML), and a Natural Sciences and Engineering Research Council Canada Discovery Grant (JM). ML’s work is carried out when she is a visiting scholar at Montclair State University.

## Notes

### Competing Interest Statement

The authors have declared no competing interest.

